# Neutralizing Activities against the Omicron Variant after a Heterologous Booster in Healthy Adults Receiving Two Doses of CoronaVac Vaccination

**DOI:** 10.1101/2022.01.28.22269986

**Authors:** Suvichada Assawakosri, Sitthichai Kanokudom, Nungruthai Suntronwong, Chompoonut Auphimai, Pornjarim Nilyanimit, Preeyaporn Vichaiwattana, Thanunrat Thongmee, Thaneeya Duangchinda, Warangkana Chantima, Pattarakul Pakchotanon, Donchida Srimuan, Thaksaporn Thatsanatorn, Sirapa Klinfueng, Ritthideach Yorsang, Natthinee Sudhinaraset, Nasamon Wanlapakorn, Juthathip Mongkolsapaya, Sittisak Honsawek, Yong Poovorawan

**Author notes:** **Corresponding author contact information** 1. Prof. Dr. Yong Poovorawan, Center of Excellence in Clinical Virology, Department of Pediatrics, Faculty of Medicine, Chulalongkorn University. Bangkok, 10330 Thailand. 2. Prof. Dr. Sittisak Honsawek, Department of Biochemistry, Osteoarthritis and Musculoskeleton Research Unit, Faculty of Medicine, Chulalongkorn University, King Chulalongkorn Memorial Hospital, Thai Red Cross Society, Bangkok, 10330, Thailand.

## Abstract

**Background:** The use of an inactivated severe acute respiratory syndrome coronavirus 2 (SARS-CoV-2) vaccine (CoronaVac) against SARS-CoV-2 is implemented worldwide. However, waning immunity and breakthrough infections have been observed. Therefore, we hypothesized that the heterologous booster might improve the protection against the delta and omicron variants.

**Methods:** A total of 224 individuals who completed the two-dose CoronaVac for six months were included. We studied reactogenicity and immunogenicity following a heterologous booster with the inactivated vaccine (BBIBP), the viral vector vaccine (AZD1222), and the mRNA vaccine (both BNT162B2 and mRNA-1273). We also determined immunogenicity at 3- and 6-months boosting intervals.

**Results:** The solicited adverse events (AEs) were mild to moderate and well-tolerated. Total RBD immunoglobulin (Ig), anti-RBD IgG, focus reduction neutralization test (FRNT50) against delta and omicron variants, and T cell response were highest in the mRNA-1273 group followed by the BNT162b2, AZD1222 and BBIBP groups, respectively. We also witnessed a higher total Ig anti-RBD in the long-interval than in the short-interval groups.

**Conclusions:** All four booster vaccines significantly increased binding and NAbs in individuals immunized with two doses of CoronaVac. The present evidence may benefit vaccine strategies development to thwart variants of concern, including the omicron variant.

## Introduction

As of January 2022, coronavirus disease 2019 (COVID-19) has spread across 200 countries with 318 million confirmed cases, and over 5.5 million deaths worldwide [1]. Vaccination is considered one of the best tools to prevent the spread of COVID-19. Several vaccines, including inactivated virus, viral vectored, mRNA, and recombinant protein subunit vaccine platforms, have been approved for emergency use against SARS-CoV-2 infection by the World Health Organization [2]. The inactivated vaccine formulation accounting for half of the 7.3 billion doses administered worldwide [3]. Due to the shortage of vaccines in Thailand, when the first vaccination campaign started in February 2021, the CoronaVac was the only effective vaccine available. However, previous studies suggested that the CoronaVac vaccine had variable efficacy in preventing severe COVID-19 infections [4,5]. Furthermore, there are several reports of waning immunity. For example, median neutralizing antibodies (NAbs) declined to near or below the detection limit after 5–8 months of CoronaVac vaccination [6–8]. Furthermore, Goldberg et al. [9] documented that the rate of breakthrough infections increased as a function of time after two doses of BNT162b2. These findings demonstrated a waning immunity after a few months of vaccination could potentially cause breakthrough infection.

During the pandemic, numerous SARS-CoV-2 variants have emerged, some of which show a high-rate of transmission and reduced effectiveness of vaccines [10]. Of these, the delta variant (B.1.617.2) has become the predominant variant, accounting for 99% of COVID-19 resurgence worldwide. The delta variant has specific mutations that have driven the explosive spread and generated immune escape [11–13]. Moreover, the delta variant is less susceptible to NAbs produced by two doses of CoronaVac, AZD1222, or BNT162b2 [14, 15]. Recently, the omicron variant (B.1.1.529) emerged in South Africa in November 2021 with several novel mutations in its receptor binding domain (RBD). Several mutations of the omicron variant escape the majority of NAbs subsets [16], which increases the transmission rate beyond that of the delta variant [17]. The immune evasion of the omicron variant has raised concerns about the protection of the vaccine against infection and development of severe disease. Although most of the population has been vaccinated, two doses of CoronaVac may not adequately protect against delta or omicron variants [18]. Therefore, additional vaccines beyond the two-dose CoronaVac regimen should be considered.

Although NAbs levels decline over time, the CD4^+^ and CD8^+^ T-cells in BNT162b2 vaccinated individuals remain and can elicit vigorous immune responses against the SARS-CoV-2 variants. Moreover, T-cells activity is minimally affected by mutations and is essential to prevent infection [19,20]. Accordingly, a previous study comparing Ad26.COV2.S and BNT162b2 as a booster dose demonstrated that the heterologous boost achieved significant increases in T cell responses than the homologous boost [21]. Furthermore, COMCOV and COV-BOOST trials in the United Kingdom (UK), demonstrated that a heterologous booster produced a higher level of immunogenicity than a homologous schedule [22,23]. Thus, a heterologous booster using a viral vector or mRNA vaccine could represent a new booster strategy and protect against infection from delta and omicron variants.

In our previous study, strong antibody response was observed after the early heterologous booster given at 3-month interval following a primary series of CoronaVac vaccination (24). Nonetheless, limited data are available on the immune response against new emerging omicron variant and how the extended time interval between primary and booster COVID-19 vaccines affected immune response. Accordingly, our study was designed to examine the reactogenicity and immunogenicity, particularly omicron variant, of four heterologous boosters given at 6-month interval. We further determined the effect of dosing interval on immune response by comparing the long-(6 months) with the short-interval (3 months). The advancement of scientific knowledge in COVID-19 booster vaccination approach including the choice of vaccine and optimal boosting interval can help overcome the waning immunity. Furthermore, booster vaccination could prevent breakthrough infection, including the newly emerged omicron variant, and support the development of new public health policies in the future.

## MATERIALS AND METHODS

### Study design and participants

This study was designed to be a prospective cohort study. Overall, 224 Thai adults aged 18–70 years old who completed the two-dose vaccination protocols for the COVID-19 inactivated vaccine or CoronaVac (Sinovac Biotech co., Ltd. Beijing, China) for 6±1 months (long-interval group) with no history of infection or diagnosed of COVID-19 were enrolled between September and December 2021. The study flow is shown in Supplement Figure 1. The exclusion criteria included any underlying diseases that may potentially induce adverse effects (AEs) after the vaccination, allergy to vaccine ingredients or vulnerable subjects.

Regarding to our previous study, between July and September 2021, 177 participants were enrolled to study safety and immunogenicity in heterologous boosters after 3±1 months of CoronaVac vaccination as published elsewhere [24]. Therefore, this group was defined as a comparison group (short-interval group). The study protocol was approved by the Institutional Review Board of the Faculty of Medicine of Chulalongkorn University (IRB numbers 546/64) and was performed according to the principles of the Declaration of Helsinki and Good Clinical Practice Guidelines (ICH-GCP). The study was registered with the Thai Clinical Trials Registry (TCTR 20210910002). Informed consent was obtained prior to participant enrolment.

### Study interventions

Participants were separated into four groups by conveniently sampling of 50–60 participants each and were assigned to receive one dose of the following vaccines: inactivated: BBIBP (Sinopharm, Beijing, China) [25]; viral vector: AZD1222 (AstraZeneca, Oxford, UK) [26]; mRNA: BNT162b2 (Pfizer-BioNTech Inc., New York City, NY, USA) [27], and mRNA-1273 (Moderna Inc., Cambridge, MA, USA) [28].

### Reactogenicity Assessment

Participants were observed under the supervision of medical professionals after vaccination to prevent acute and severe side effects. AEs were recorded, including injection site pain, induration, redness, fever, headache, muscle pain, and others. The participants then received an AE record form for further self-monitoring within 7 days after vaccination.

### Sample collection, total RBD Ig, and anti-RBD IgG/nucleocapsid assay

Peripheral venous blood samples were collected to measure the immune response, including the total Ig against RBD, IgG against RBD, and IgG against nucleocapsid (N) as previously described [24].

### Surrogate virus neutralization assay to wild-type, alpha, beta, delta, and omicron variants

To measure NAbs against the SARS-CoV-2 variants, the cPass™ SARS-CoV-2 surrogate virus neutralization test Kit was used (GenScript Biotech, New Jersey, USA), consisting of recombinant RBD from alpha, beta, delta, and omicron variants as previously described [24].

### Focus reduction neutralization test

The neutralization ability of antibodies in vaccinated serum was measured using the FRNT50. Briefly, serum was heated for 30 min at 56°C and then diluted serially (1:10–1:7,290). The serum was mixed with the SARS-CoV-2 variant (delta and omicron) and incubated for 1 hour at 37°C. The mixtures were then transferred to a 96-well plate containing confluent Vero cell monolayers and incubated for 2 hours, then 1.5% semi-solid carboxymethyl cellulose was added to limit virus diffusion and cells were incubated further at 37°C, 5% CO2. Next, Vero cells were fixed with 3.7% formaldehyde/PBS and permeabilized with 2% Triton X-100. Anti-NP human mAb was then added, followed by peroxidase-conjugated goat anti-human IgG. Finally, the infected cells were visualized after adding TrueBlue Peroxidase Substrate. The foci of virus infected cells were counted by CTL ImmunoSpot S6 analyzer. The percentage of focus reduction was calculated and IC50 was determined by PROBIT software. The detection limit is 1:20 [29].

### Interferon-gamma release assay

QuantiFERON (QFN) SARS-CoV-2 RUO (Qiagen, Hilden, Germany) interferon-gamma (IFN-γ) release assay was performed to measure T cell response as previously described [24]. The seropositivity rate was calculated using IFN-γ levels from the stimulated tube minus the negative control tube. An IFN-γ level above 0.15 IU/mL was defined as positive.

### Statistical analysis

All statistical analyses were conducted using GraphPad Prism v9.0 (GraphPad Software, San Diego, CA, USA), the Statistical package for the social sciences (SPSS) v.22 (SPSS Inc., Chicago, IL, USA), and R v4.1.2. Software (R Foundation for Statistical Computing, Vienna, Austria). The normality of the data was tested using the Kolmogorov–Smirnov test. Statistical comparison between qualitative data was performed using Pearson’s χ^2^ or Fisher’s exact test. Comparison between quantitative data was performed using ANOVA or the Kruskal–Wallis H test, where appropriate. The geometric means of the antibody titers and confidence intervals were determined using R. the Wilcoxon signed-rank test was used for pairwise analysis. A *p*-value <0.05 was considered statistically significant.

## RESULTS

### Demographic data

Between September 2021 and December 2021, 224 participants were eligible to be included in the study. All participants were healthy Thai adults aged 18 years or older who had received two doses of CoronaVac for 6±1 months from the first dose at the time of enrollment. Among all enrolled participants, 117 (52.2%) were female and 107 (47.8%) were male. The mean age (range) of the participants in the BBIBP-CorV, AZD1222, BNT162b2, and mRNA-1273 groups was 41.91 (24–64), 44.13 (21–59), 41.56 (22–58), and 37.05 (21–57), respectively. Common underlying diseases included allergy, diabetes mellitus, dyslipidemia, and hypertension. The follow-up interval between visit 1/2 and visit 1/3 was 14.66 (13–20) and 28.27 (22–35) days. The baseline characteristics in four groups of participants were similar (Table 1). Two participants were excluded after testing positive for SARS-CoV-2 infection, and two participants were lost to follow-up during the study. All participants provided written informed consent.

**Table 1.**
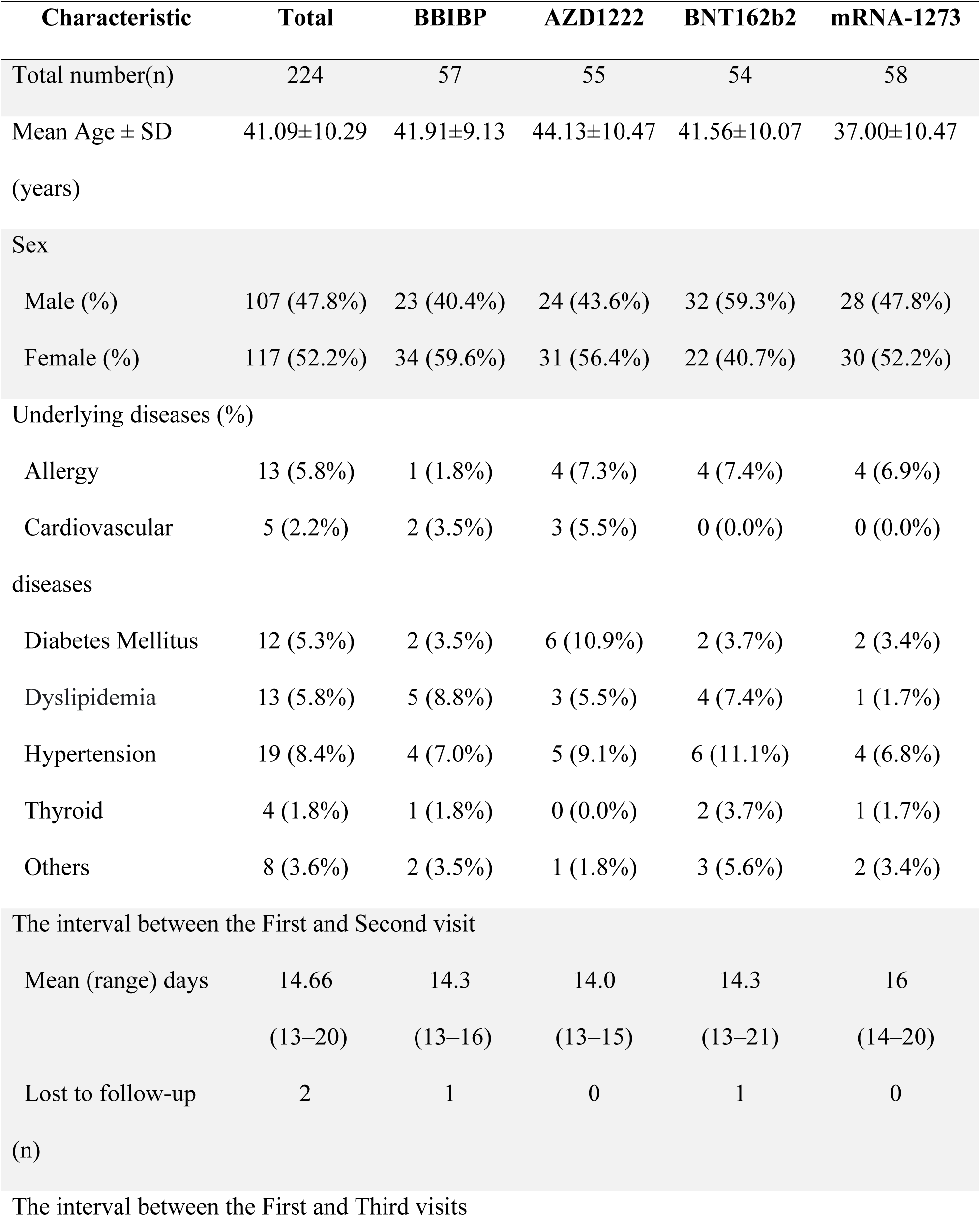
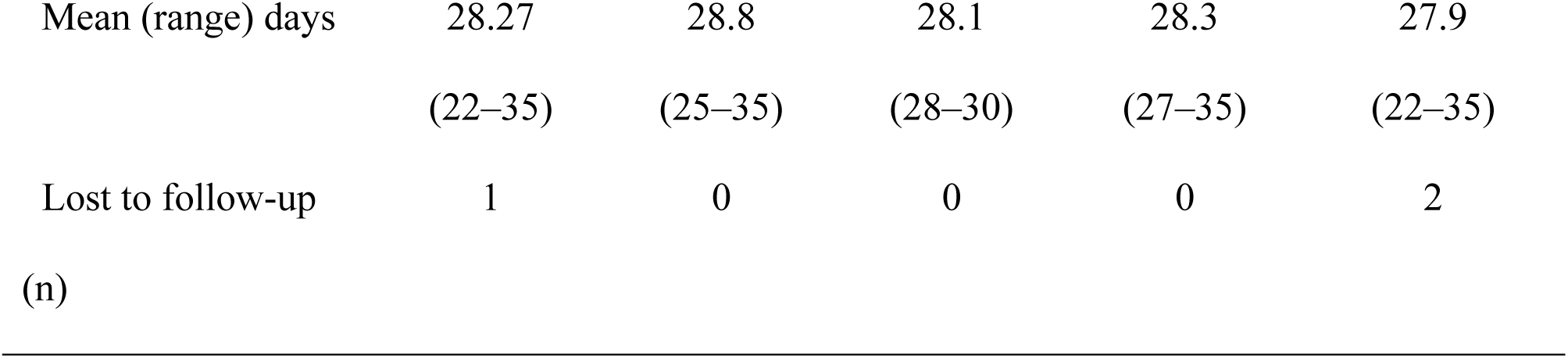
Baseline characteristics of participants enrolled in this study.

### Reactogenicity data

The AE profile, including dizziness, nausea, vomiting, and diarrhea, was similar within 7 days after vaccination. Generally, the severity of local and systemic reactions was reported as mild or moderate and resolved within a few days post-vaccination. The most observed local AE was injection site pain, accounting for 34 (59.6%), 46 (83.6%), 52 (96.3%), and 56 (94.9%) and the most observed systemic AE was myalgia, accounting for 24 (42.1%), 41 (74.5%), 38 (70.4%), and 46 (78.0%) in the BBIBP, AZD1222, BNT162b2, and mRNA-1273 groups, respectively (Figure 1A–1D). The analysis of AEs revealed that pain at the injection site, swelling, redness, myalgia, joint pain, and chilling in individuals who received BBIBP were significantly less frequent than the other vaccine platforms (Supplementary Figure 2A–2C). In comparison, systemic reactions such as fever, headache, chilling, and joint pain were highly evidenced after receiving AZD1222 and mRNA-1273. No serious AEs were reported during the study (Supplementary Table 1).

**Figure 1.**
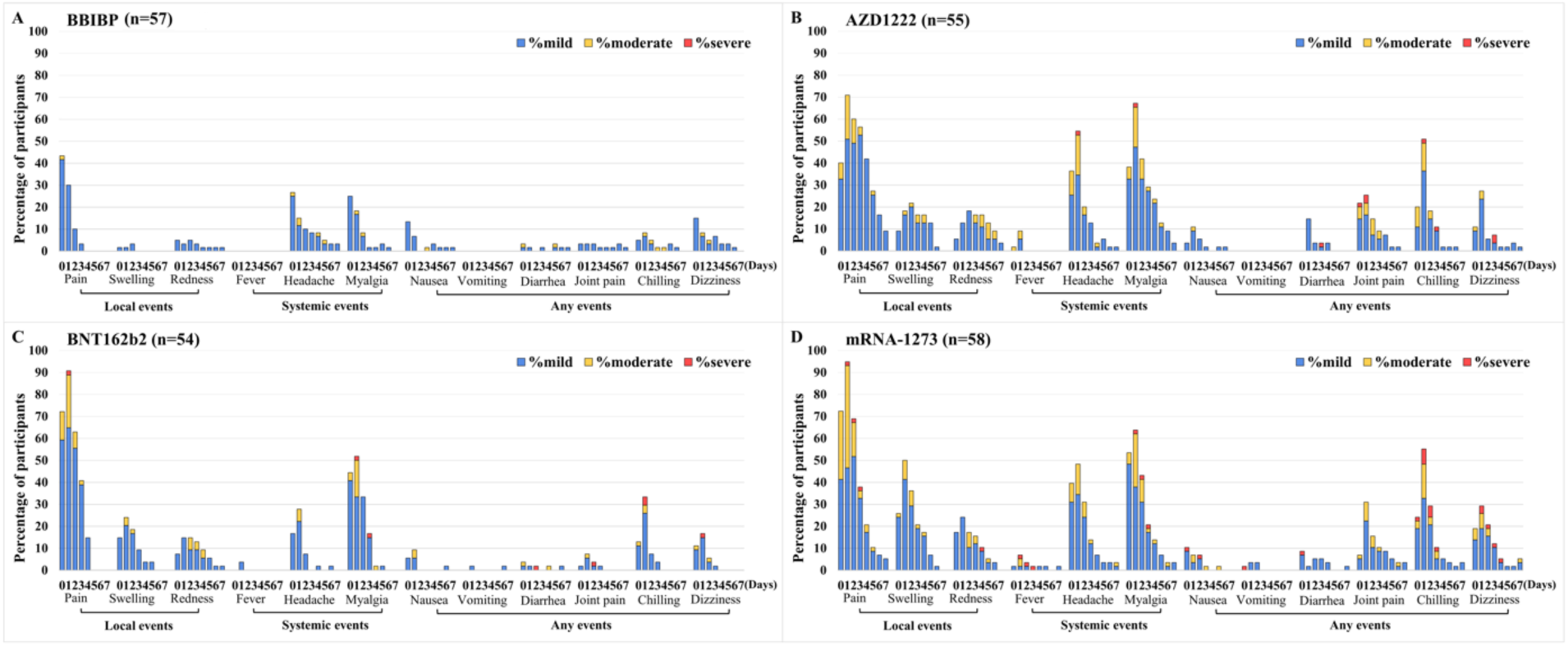
The reactogenicity and severity solicited local and systemic reactions over the first 7 days following vaccination with a booster dose of COVID-19 vaccines. Booster vaccine included the inactivated vaccine – BBIBP **(Panel A)**, viral vector vaccine – AZD1222 **(Panel B)**, mRNA vaccine – BNT162b2 **(Panel C)**, and mRNA vaccine –mRNA-1273 **(Panel D)**. The percentage of participants who report adverse events were presented on the Y-axis. The swelling and redness were graded by measuring the diameter area as mild (<5 cm), moderate (5 to <10 cm), and severe (≥10 cm). Fever was graded as mild (38°C to <38.5°C), moderate (38.5°C to <39°C), and severe (≥39°C). The other events were graded as mild (no limitation on regular activity)/Moderate (some limitation of daily activity)/severe (unable to perform or prevented daily activity).

### Humoral immune responses after booster dose vaccination at 6 months

At the baseline visit, all participants presented seropositivity for total RBD Ig and anti-RBD IgG. There were no significant differences in the geometric mean titer (GMT) of the total RBD Ig among all groups at baseline. Interestingly, the COVID-19 vaccines administered as the booster dose significantly elicited higher total RBD Ig levels at 14 days post-vaccination, with GMTs of 1740, 12260, 31793, and 51979 U/mL (*p<*0.001). Subsequently, the total RBD Ig was slightly reduced to 1295, 12111, 21053, and 33519 U/mL (*p<*0.001) in the BBIBP, AZD1222, BNT162b2, and mRNA-1273 groups, at 28 days post-vaccination, respectively (Figure 2A). Comparable trends were observed with anti-RBD IgG levels (Figure 2B). Following the administration of the third booster, the mRNA-1273-boosted individuals possessed the highest total Ig and anti-RBD IgG levels. However, there was no significant difference in the responses between the two mRNA vaccines. After boosting with BBIBP, the total RBD Ig and anti-RBD IgG levels were significantly lower than the levels achieved by viral-vector and mRNA groups (Supplementary Table 2). Anti-N IgG levels were also measured at three different time points, as shown in Figure 2C. At baseline, the median level (IQR) of anti-N IgG was 0.13 (0.07–0.31). The results were comparable among the four groups, which could be interpreted that none of 224 participants had been infected by SARS-CoV-2. After the booster dose, the median anti-N IgG level increased significantly to 2.78 (1.84-5.30) in those who received BBIBP (*p<*0.001). On the contrary, no change in anti-N IgG levels was observed after the AZD1222 or mRNA vaccination because only the inactivated vaccine contained the SARS-CoV-2 N protein in the formulation (Figure 2C).

**Figure 2.**
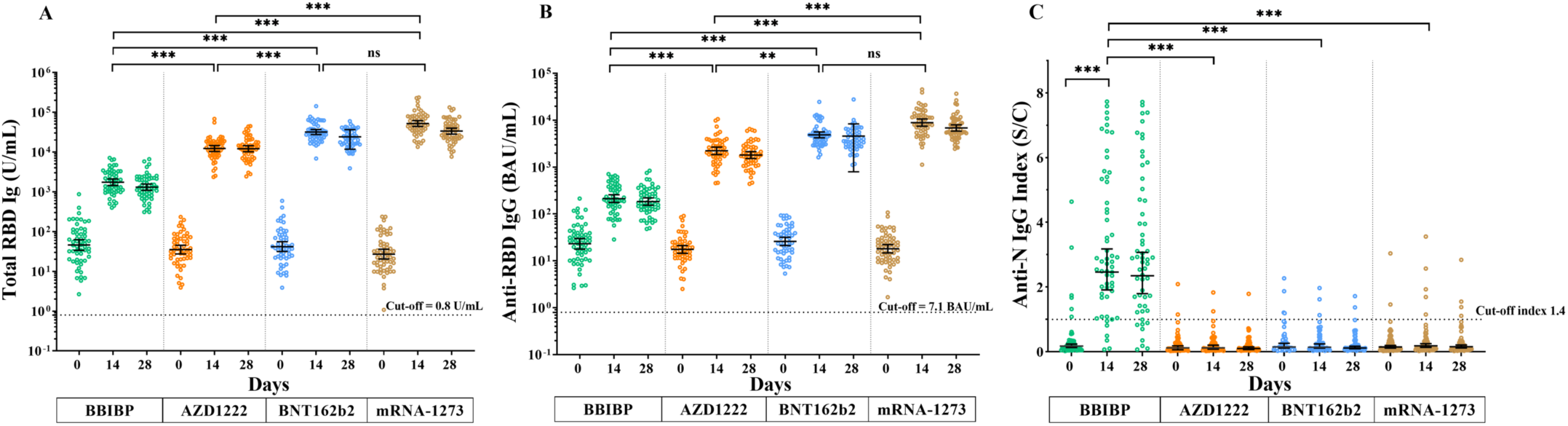
Detection of SARS-CoV-2 specific humoral responses. **Panel A** shows the total RBD Ig of SARS-CoV-2 (U/mL). **Panel B** shows the anti-RBD IgG of SARS-CoV-2 (BAU/mL) and **Panel C** anti-N IgG of SARS-CoV-2 index (S/C). Each data point represents an individual who completed two doses of CoronaVac, followed by the inactivated vaccine, BBIBP (green), the viral-vector vaccine, AZD1222 (orange), the mRNA vaccine, BNT162b2 (blue), or mRNA-1273 (gold). Lines indicate GMT, and the I bar indicates 95% confidence intervals (95% CI). ns indicates no statistical difference; ***, *p*<0.001.

### Surrogate virus neutralization specific to wild-type and variants of concern

The surrogate virus neutralization assay was performed to compare the functionality of NAbs specific to the wild-type and SARS-CoV-2 variant strains, including alpha, beta, delta, and omicron. Overall, the seropositivity rate of individuals who received two doses of CoronaVac was 6/40 (15%), 5/40 (12.5%), 1/40 (2.5%), 8/40 (20%), and 0/40 (0%) to wild-type, alpha, beta, delta, and omicron variants, respectively (Figure 3A–3D and Supplementary Figure 3). Sera samples from individuals vaccinated with AZD1222, BNT162b2, and mRNA-1273 showed higher than 97% neutralizing activity against wild-type and SARS-CoV-2 variants except for the omicron variants (Supplementary Table 3). For the omicron variant, seropositivity rate was detected in 2/30 (6.7%) BBIBP, 19/30 (63.3%) AZD1222, 21/30 (70.0%) BNT162b2, and 27/30 (90.0%) mRNA-1273 vaccinated individuals (Figure 3A–3D). These data suggested that breadth of neutralizing activities against the omicron variant were reduced in all vaccinated groups compared to those against other VOCs.

**Figure 3.**
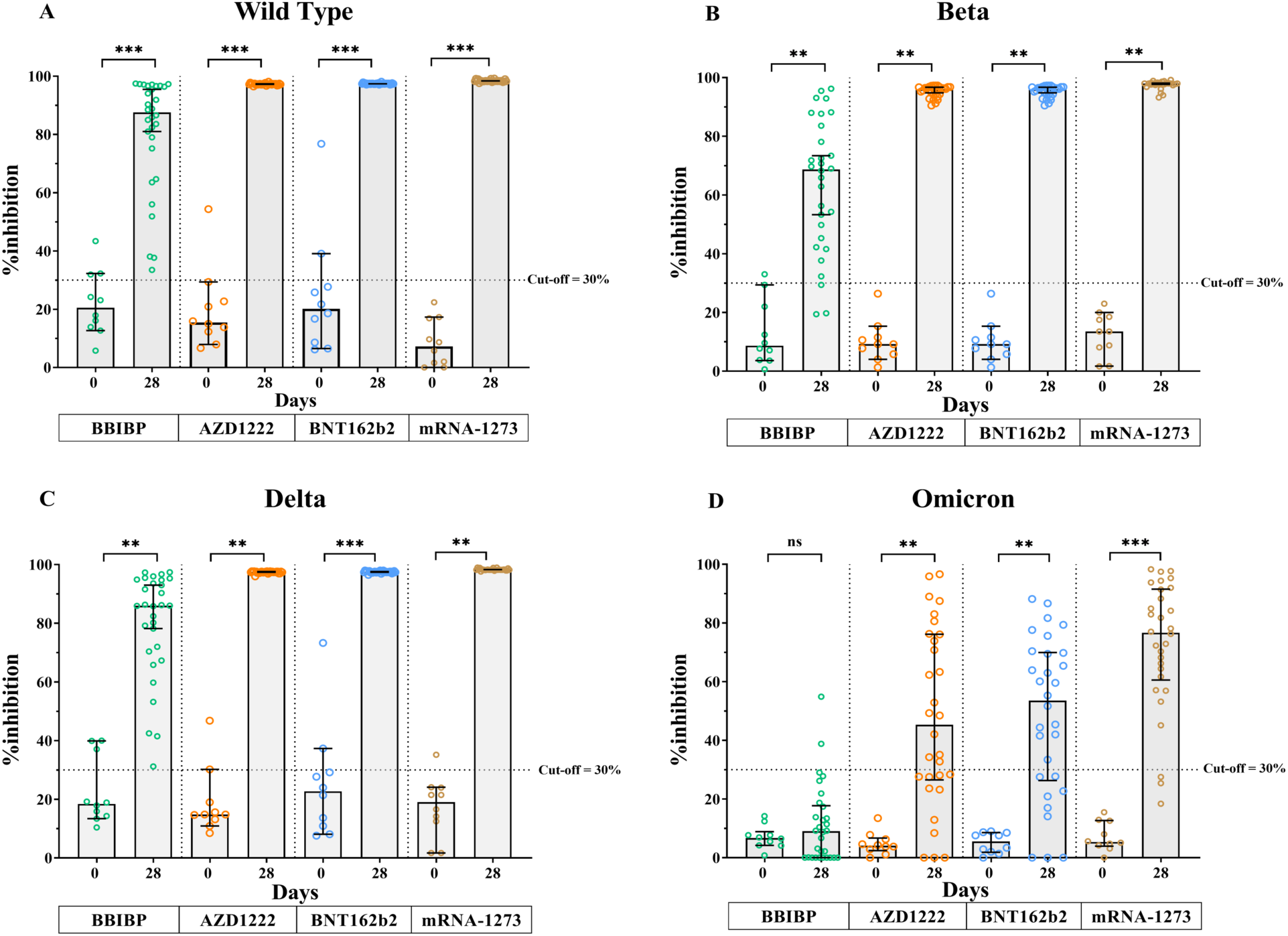
The neutralizing activities using a surrogate virus neutralization test. The neutralizing activities specific to SARS-CoV-2 wild-type (**Panel A**) and the variant of concerns including beta (**Panel B**), delta (**Panel C**), and omicron (**Panel D**) were shown as %inhibition. Each data point represents an individual who received a heterologous third booster vaccine, the inactivated vaccine, BBIBP (green), the viral-vector vaccine, AZD1222 (orange), the mRNA vaccine, BNT162b2 (blue), or mRNA-1273 (gold). Lines represent median (IQR). ns indicates no significant difference; *\*\*, p*<0.01; ***, *p*<0.001.

### Focus reduction neutralization test

The NAbs titers in sera from participants were determined using a live virus neutralization assay, FRNT50. For all participants, undetectable (titer<20) NAbs at baseline were observed in 119/120 and 114/120 to the delta and omicron variants, respectively. The GMTs (range) against the delta variant were 69.6 (44.8–108), 1003 (744–1352), 1285 (982–1680), and 2168 (1627– 2889) at 28 days after receiving a booster dose with BBIBP, AZD1222, BNT162b2, and mRNA-1273 vaccines, respectively (Figure 4A). Lower GMT values were observed against the omicron variant: 24.6 (18.1–33.5), 250 (169–368), 277 (190–402), and 512 (359–732) (Figure 4B). Compared to the delta variant, the FRNT 50 values against omicron were reduced 2.8-fold for BBIBP, 4.0-fold for AZD1222, 4.6-fold for BNT162b2, and 4.2-fold for mRNA-1273.

**Figure 4.**
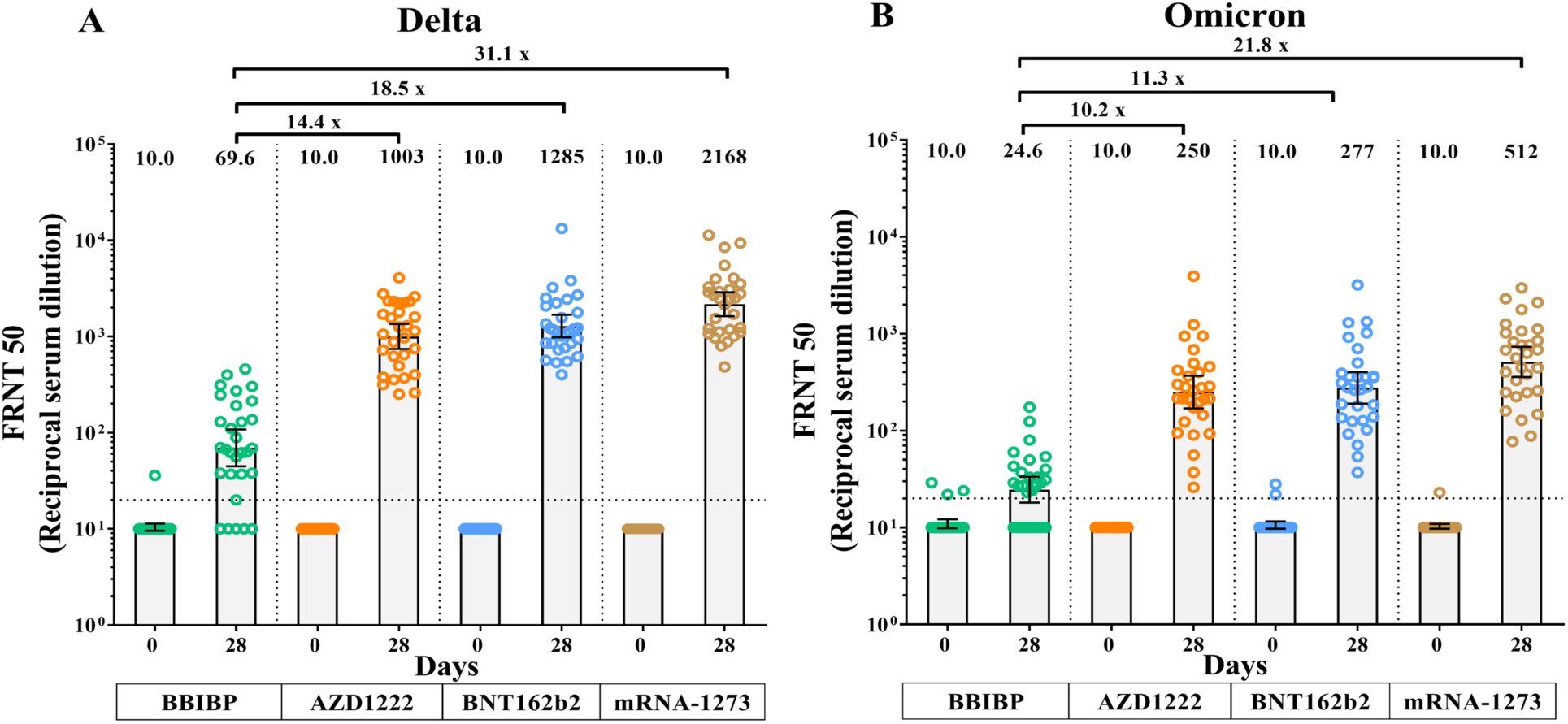
Neutralizing antibody titres of SARS-CoV-2 before and 28 days after boost vaccination for delta and omicron variants. Neutralization of SARS-CoV-2 was measured using the focus reduction neutralization test (FRNT). **Panel A** shows the neutralization against the delta variant, and **Panel B** shows the neutralization against the omicron variant. Each data point represents an individual who received a heterologous third booster vaccine, the inactivated vaccine, BBIBP (green), the viral-vector vaccine, AZD1222 (orange), the mRNA vaccine, BNT162b2 (blue), or mRNA-1273 (gold). Error bars show GMT and 95% CI. Values below the limit were substituted with a titer of 10.

### Total interferon-gamma response following immunization

In addition to the serological responses, we evaluated the presence of T cell responses by measuring the total IFN-γ level using the QFN SARS-CoV-2-ELISA assay. The seropositivity rates for the IFN-γ CD4+/IFN-γ CD4+ and CD8+ levels observed in most participants after the third booster with 86%/93% for AZD1222, 96%/100% for BNT162b2, and 90%/93% for mRNA-1273 (Figure 5A and 5B). In contrast, the seropositivity rate was 43%/47% for BBIBP, suggesting a lower T-cell response was elicited. Consistent with the total RBD Ig levels, the T-cell-related IFN-γ response was significantly elevated at 14 days compared to baseline and gradually declined at 28 days after vaccination (Supplementary Table 4). Our result suggests that a single booster dose with AZD1222, mRNA-1273, or BNT162b2 could rapidly induce a T-cell response in individuals vaccinated with CoronaVac.

**Figure 5.**
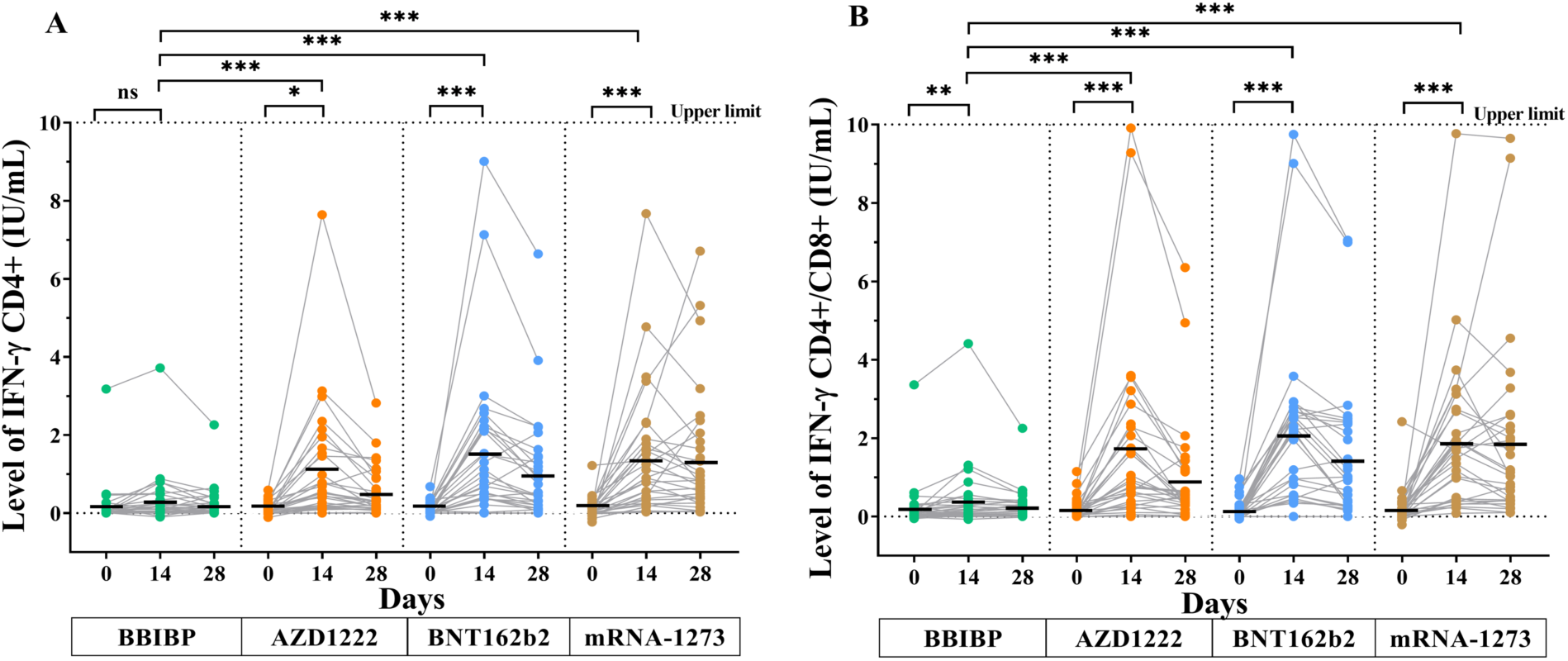
Interferon-gamma release assay. **Panel A** shows the IFN-γ produced by CD4+ T cell, and **Panel B** shows the IFN-γ produced by CD4+ and CD8+ T cell. QFN IFN-γ ELISA was used to evaluate the IFN-γ levels at 14 days and 28 days following the third booster of inactivated vaccine, BBIBP (green), the viral-vector vaccine, AZD1222 (Orange), or the mRNA vaccine, BNT162b2 (blue), and mRNA-1273 (gold). Horizontal bars indicate median of the IFN-γ levels. ns indicates no significant difference; *, *p*<0.05, **, *p*<0.01, ***, *p*<0.001.

### Comparison of total RBD Ig levels between short-and long-interval groups

We compared GMT levels of total RBD Ig between short-and long-interval groups. As shown in Figure 6A–6C, the long-interval group elicited higher total RBD Ig than the short-interval group at 14 days post-booster vaccination. Compared to the pre-booster level, the BBIBP vaccine significantly induced a 30.2-fold increase in total RBD Ig titer in the short-interval group and a 37.8-fold increase in the long-interval group. In contrast, AZD1222 and BNT162b2 induced 209.1-fold and 387.0-fold for the short-interval group and 307.5-fold and 674.7-fold for the long-interval group, respectively.

**Figure 6.**
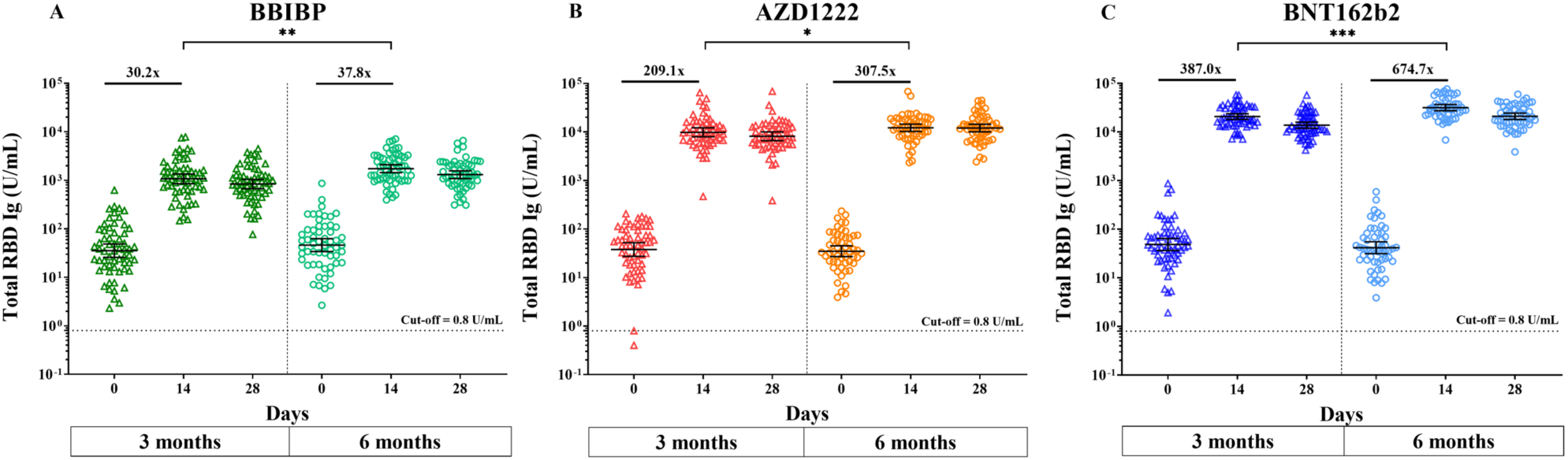
Compare the total RBD Ig (U/mL) between the short interval (triangle) and the long interval (circle). The total RBD Ig (U/mL) at 14 days following the third booster of inactivated vaccine, BBIBP **(Panel A)**, the viral-vector vaccine, AZD1222 **(Panel B)**, or the mRNA vaccine, and BNT162b2 **(Panel C)**. ns indicates no significant difference; *, *p*<0.05, **, *p*<0.01, ***, *p*<0.001 (***).

## DISCUSSION

We evaluated the immunogenicity and safety profile following the administration of heterologous vaccines as third boosters in individuals previously vaccinated with two doses of CoronaVac vaccines for 6 months. The magnitude of the total Ig anti-RBD (U/mL) and IgG-specific RBD (BAU/mL) responses was higher in individuals receiving the viral vector vaccines and mRNA vaccine than the inactivated vaccine. In addition, a substantial increase in the neutralizing potency against wild type and SARS-CoV-2 variants was observed in individuals who received a third heterologous booster compared to baseline. In comparing the third heterologous booster, the mRNA and viral vectored vaccines elicited neutralizing titers with a 14–31-fold increase for delta and a 10–22-fold increase for omicron compared to individuals receiving the inactivated vaccine. Regarding the reactogenicity after the third dose, all vaccines have acceptable AEs. The results showed that mild to moderate local and systemic side-effects were observed more frequent in individuals receiving mRNA or viral vector than in those receiving inactivated platforms within the first seven days of vaccination, which was similar to the findings observed in the previous study of third heterologous boosters, primed with two AZD1222 or BNT162b2 vaccines [23]. These indicated that the AEs were tolerable after boosters with different vaccines. In conclusion, no serious AEs were reported among the groups, indicating that the heterologous third booster dose has a good safety profile.

Consistent with a previous study of CoronaVac, we observed that the baseline total IgG levels were reduced at six months after the primary series of CoronaVac [8]. The data indicated that the immunogenicity from CoronaVac is waning over time. As expected, the heterologous booster with BBIBP, AZD1222, BNT162b2, or mRNA-1273 significantly elicited an immune response at 14 days compared to the baseline level. Our data suggested that a single-shot booster after a two-dose of CoronaVac may be sufficient to restore the B cell memory response [30]. Regarding the total Ig RBD and anti-RBD IgG response, some differences were observed among the four boosters. Following the administration of the inactivated vaccine, there was a lower antibody response post-booster similar to that observed in a previous study from Brazil [31].

The neutralization susceptibility for the omicron variant was continuously reduced by vaccine-induced immunity [32]. Boosting with heterologous vaccines might induce the memory B-cells evolved through affinity maturation, leading to a broader and more potent NAbs response against SARS-CoV-2 variants [33,34]. Our study revealed increased cross-neutralizing activities for the variant after receiving a third booster dose compared to a second dose. Similar result was found in individuals who received two doses of mRNA vaccines and were subsequently vaccinated with mRNA vaccine [35]. Nevertheless, not all heterologous boosted vaccines could elicit a substantial increase in NAbs against omicron variants. Furthermore, we found that individuals who received the mRNA vaccine as a third dose produce the highest neutralizing activities followed by a viral vectored and inactivated vaccine. Supporting studies found that heterologous mRNA booster following two doses of inactivated vaccines could elicit higher neutralizing antibodies than the inactivated vaccine boosted [36,37]

Our findings showed that the reduction of NAbs levels against omicron was much lower relative to delta variants. Consistent with a previous study, the neutralizing titer against the omicron variant was 4.9-fold less than that for the delta variant with the BBIBP-CorV/ZF2001 heterologous booster [38] and was 24.5-fold lower than that for the delta variant in sera from individuals who received BNT162b2 as the third dose following two doses of BNT162b2 [39]. The substantially reduced of neutralization against the omicron variant may be related to several mutations concentrated around the RBD in the spike protein compared to other variants [40].

Although previous studies reported a potential immune escape by SARS-CoV-2 variants at the antibody level [41], little is known at the T-cell level. Recent studies have demonstrated that most CD4+ and CD8+ T-cells could cross-recognize SARS-CoV-2 variants harboring S mutations [20,42,43]. According to our study, the CD4+ and CD8+ T cell-producing IFN-*γ* were rapidly provoked after the AZD1222 or mRNA vaccine boosters. Consistent with the magnitude of the antibody responses, the T-cell response was lower in the BBIBP group compared with the other boosters. Beyond humoral responses, this study may indicate that the booster vaccines AZD1222, BNT162b2, or mRNA-1273 could contribute to cross-protection against SARS-CoV-2 variants by enhancing the adaptive T-cells immune response [44].

Moreover, a long interval between the primary and the boosters could significantly stimulate a better immune response than a shorter interval. A similar previous study using the extended interval also suggested that antibody responses after receiving the third booster AZD1222 and BNT162b2 vaccine were higher in extended interval vaccinations [45]. In summary, these data indicated that a longer interval might allow more time for enhancing immune memory responses.

An early estimate of vaccine effectiveness (VE) in Chile, the study found that the adjusted VE against symptomatic, hospitalization, and COVID-19 related deaths due to the delta variant was increased after a heterologous booster shot with a viral-vectored and mRNA vaccine following a complete primary series of CoronaVac vaccines [46]. However, there were limited data in that study on the VE against omicron infection following a heterologous booster. Our results highlight that the viral vector and mRNA vaccines, but not inactivated vaccines, were promising booster vaccines that could elicit a strong immune response to prevent the omicron variant infection.

This study had some limitations. First, the study did not examine the phenotype of CD4+ and CD 8+ T cells. Moreover, the total IFN-γ level and the surrogate virus neutralization test reached the upper limit of detection assay in some samples. Thus, we could not estimate the exact cellular immune response following the third booster vaccine. Further study using more advanced techniques included activation-induced marker assays may resolve this issue. Moreover, long-term studies are needed to determine the durability of the booster vaccination.

In summary, we report a robust total RBD Ig response and acceptable safety profile after implementing heterologous third booster vaccines. However, neutralizing activities against omicron variants and the T-cell response were reduced in BBIBP-boosted individuals compared to AZD1222, BNT162b2, and mRNA 1273-boosted individuals.

## Data Availability

All data produced in the present work are contained in the manuscript

## Footnote

## Acknowledgments

We would like to thank the staff of the Center of Excellence in Clinical Virology and all the participants for helping and supporting in this project. We also thank the Ministry of Public Health, Chulabhorn Royal Academy and Zullig pharma for providing the vaccines for this study.

