## Supplementary Information for "Neutralizing Activities against the Omicron Variant after a Heterologous Booster in Healthy Adults Receiving Two Doses of CoronaVac Vaccination"

**Supplementary Table 1.** Statistical analysis of reactogenicity data between booster vaccines.

| | Total | BBIBP | AZD1222 | BNT162b2 | mRNA-1273 | Pearson's $\chi^2$ | Pair-wise analysis (Fisher's Exact test) | | | | | |
|---|---|---|---|---|---|---|---|---|---|---|---|---|
| | | | | | | | 1 vs 2 | 1 vs 3 | 1 vs 4 | 2 vs 3 | 2 vs 4 | 3 vs 4 |
| **N (%)** | 224 (100) | 57 (100) | 55 (100) | 54 (100) | 58 (100) | | | | | | | |
| **Injection site pain** | 187 (83.5) | 34 (59.6) | 46 (83.6) | 52 (96.3) | 55 (94.8) | < 0.001 | 0.006 | < 0.001 | < 0.001 | 0.052 | 0.069 | 1.000 |
| **Swelling** | 71 (31.7) | 2 (3.5) | 19 (34.5) | 16 (29.6) | 34 (58.6) | < 0.001 | < 0.001 | < 0.001 | < 0.001 | 0.683 | 0.010 | 0.002 |
| **Redness** | 63 (28.1) | 6 (13.7) | 16 (29.1) | 18 (33.3) | 23 (39.7) | 0.004 | 0.017 | 0.005 | < 0.001 | 0.683 | 0.322 | 0.558 |
| **Fever** | 11 (4.9) | 0 (0.0) | 5 (9.1) | 2 (3.7) | 4 (6.9) | 0.046 [a] | 0.026 | 0.234 | 0.119 | 0.438 | 0.738 | 0.680 |
| **Headache** | 120 (53.6) | 28 (49.1) | 34 (61.8) | 20 (37.0) | 38 (65.5) | 0.011 | 0.189 | 0.251 | 0.091 | 0.013 | 0.700 | 0.004 |
| **Myalgia** | 148 (66.1) | 24 (42.1) | 41 (74.5) | 38 (70.4) | 45 (77.6) | < 0.001 | 0.001 | 0.004 | < 0.001 | 0.672 | 0.826 | 0.398 |
| **Nausea** | 37 (16.5) | 9 (15.8) | 11 (20.0) | 7 (13.0) | 10 (17.2) | 0.796 | 0.627 | 0.789 | 1.000 | 0.440 | 0.810 | 0.604 |
| **Vomiting** | 6 (2.7) | 0 (0.0) | 0 (0.0) | 2 (3.7) | 4 (6.9) | 0.029 [a] | N/A | 0.234 | 0.119 | 0.243 | 0.119 | 0.680 |
| **Diarrhoea** | 32 (14.3) | 6 (10.5) | 12 (21.8) | 4 (7.4) | 10 (17.2) | 0.127 | 0.127 | 0.743 | 0.420 | 0.056 | 0.637 | 0.155 |
| **Joint pain** | 53 (23.7) | 7 (12.3) | 19 (34.5) | 7 (13.0) | 20 (34.5) | 0.002 | 0.007 | 1.000 | 0.008 | 0.013 | 1.000 | 0.009 |
| **Chilling** | 101 (45.1) | 9 (15.8) | 32 (58.2) | 20 (37.0) | 40 (69.0) | < 0.001 | < 0.001 | 0.017 | < 0.001 | 0.035 | 0.248 | 0.001 |
| **Dizziness** | 67 (39.9) | 13 (22.8) | 17 (30.9) | 14 (25.9) | 23 (49.7) | 0.218 | 0.396 | 0.825 | 0.070 | 0.672 | 0.431 | 0.160 |

[a] Likelihood Ratio is applied due to expected cell count less than 5 more than 20% of cells

**Supplementary Table 2.** The value and statistical analysis of GMT (with 95% confidence intervals) of total RBD Ig and anti-RBD IgG, median (with interquartile ranges) of anti-N IgG.

| | BBIBP (I) | AZD1222 (II) | BNT162b2 (III) | mRNA-1273 (IV) | Total | Kruskal-Wallis | Pair-wise analysis (Mann Whitney) | | | | | |
|---|---|---|---|---|---|---|---|---|---|---|---|---|
| | | | | | | | (I) vs (II) | (I) vs (III) | (I) vs (IV) | (II) vs (III) | (II) vs (IV) | (III) vs (IV) |
| **SARS-CoV-2 total RBD Ig, (U/ml), GMT 95% CI]** | | | | | | | | | | | | |
| **Day 0** | 46.01 (33.87-62.50) | 35.17 (27.32-45.26) | 41.67 (31.38-55.32) | 27.07 (20.44-35.84) | 36.49 (31.72-41.97) | 0.033 [a] | 1.000 | 1.000 | 0.035 | 1.000 | 1.000 [b] | 0.156 |
| **Day 14** | 1740.00 (1443.81 -2096.95) | 12259.51 (10333.78 -14544.10) | 31792.88 (27428.26 -36852.03) | 51979.58 (43920.69-61516.02) | 13650.90 (11269.99-16534.79) | < 0.001 | < 0.001 | < 0.001 | < 0.001 | < 0.001 | < 0.001[b] | 0.099 |
| **Day 28** | 1294.54 (1084.08 -1545.87) | 12111.12 (10153.22 -14446.57) | 21053.33 (18145.07 -24427.73) | 33519.20 (28228.32-39802.45) | 10146.77 (8426.09-12218.84) | < 0.001 | < 0.001 | < 0.001 | < 0.001 | 0.017 | < 0.001 [b] | 0.085 |
| **SARS-CoV-2 anti-RBD IgG, (BAU/ml), GMT [95% CI]** | | | | | | | | | | | | |
| **Day 0** | 23.16 (17.83-30.08) | 17.55 (14.52-21.23) | 26.01 (21.16-31.97) | 18.13 (14.78-22.23) | 20.85 (18.71-23.23) | 0.028 [a] | 0.435 | 1.000 | 0.609 | 0.073 | 1.000 [b] | 0.109 |
| **Day 14** | 212.55 (175.84 -256.93) | 2226.68 (1858.02 -2668.49) | 4877.73 (4221.82 -5635.55) | 8901.15 (7471.96-10605.14) | 2120.51 (1723.96-2608.29) | < 0.001 | < 0.001 | < 0.001 | < 0.001 | 0.001 | < 0.001 [b] | 0.017 |
| **Day 28** | 183.83 (153.94 -219.51) | 1808.49 (1528.28 -2140.07) | 3836.95 (3292.30 -4471.70) | 6846.00 (5867.92-7987.17) | 1702.33 (1393.57-2079.51) | < 0.001 | < 0.001 | < 0.001 | < 0.001 | 0.002 | < 0.001 [b] | 0.012 |
| **SARS-CoV-2 anti-N IgG, (S/C), median [IQR]** | | | | | | | | | | | | |
| **Day 0** | 0.16 (0.08-0.32) | 0.12 (0.05-0.26) | 0.15 (0.10-0.36) | 0.13 (0.07-0.33) | 0.13 (0.07-0.31) | 0.251 | ns | ns | ns | ns | ns | ns |
| **Day 14** | 2.78 (1.84-5.30) | 0.12 (0.05-0.27) | 0.14 (0.09-0.41) | 0.14 (0.09-0.37) | 0.24 (0.09-1.13) | < 0.001 | < 0.001 | < 0.001 | < 0.001 | 1.000 | 1.000 | 1.000 |
| **Day 28** | 2.89 (1.55-4.87) | 0.10 (0.06-0.23) | 0.12 (0.08-0.31) | 0.12 (0.07-0.31) | 0.19 (0.08-1.02) | < 0.001 | < 0.001 | < 0.001 | < 0.001 | 1.000 | 1.000 [b] | 1.000 |

[a] tested using student's t-test; tests were adjusted by Bonferroni correction for multiple tests.

[b] adjusted for age

**Supplementary Table 3.** The percentage inhibition against wild-type and SARS-CoV-2 variants.

| | BBIBP (I) | AZD1222 (II) | BNT162b2 (III) | mRNA-1273 (IV) |
|---|---|---|---|---|
| **sVNT—Wild Type. (GenScript), median (IQR)** | | | | |
| Day 0 | 20.50 (13.60-32.00) | 15.45 (11.20-24.38) | 20.15 (8.075-30.55) | 7.250 (1.20-17.00) |
| Day 28 | 87.55 (72.58-96.63) | 97.30 (97.10-97.50) | 97.50 (97.30-97.50) | 98.30 (98.30-98.40) |
| **sVNT—Alpha. (GenScript), median (IQR)** | | | | |
| Day 0 | 22.0 (14.4-29.2) | 20.6 (15.5-23.6) | 23.8 (15.8-27.0) | 5.10 (0.075-8.48) |
| Day 28 | 75.4 (55.8-92.4) | 97.9 (97.5-98.3) | 98.1 (97.9-98.3) | 98.2 (98.2-98.3) |
| **sVNT—Beta. (GenScript), median (IQR)** | | | | |
| Day 0 | 8.65 (3.75-23.9) | 9.20 (5.35-12.5) | 9.20 (5.35-12.5) | 13.5 (6.50-18.9) |
| Day 28 | 68.7 (44.5-84.6) | 96.0 (94.2-96.8) | 96.0 (94.2-96.8) | 98.0 (97.6-98.2) |
| **sVNT—Delta. (GenScript), median (IQR)** | | | | |
| Day 0 | 18.4 (14.1-37.8) | 14.8 (12.6-21.8) | 22.7 (10.2-31.2) | 19.1 (9.80-24.0) |
| Day 28 | 86.0 (69.6-94.8) | 97.5 (97.3-97.6) | 97.5 (97.3-97.6) | 98.2 (98.2-98.3) |
| **sVNT—Omicron. (GenScript), median (IQR)** | | | | |
| Day 0 | 6.70 (4.18-8.88) | 4.05 (2.45-6.68) | 5.50 (1.85 -8.55) | 5.35 (3.93-12.7) |
| Day 28 | 9.00 (0.00-17.7) | 45.3 (26.5-76.2) | 53.5 (26.3-70.0) | 76.7 (60.6-91.6) |

**Supplementary Table 4.** The IFN-γ CD4+/IFN-γ CD4+ and CD8+ levels

| | BBIBP (I) | AZD1222 (II) | BNT162b2 (III) | mRNA-1273 (IV) |
|---|---|---|---|---|
| **IFN-γ CD4+ T-cell (IU/ml), median (IQR)** | | | | |
| Day 0 | 0.04 (0.00-0.155) | 0.05 (0.01-0.22) | 0.05 (0.01-0.19) | 0.03 (0.01-0.20) |
| Day 14 | 0.09 (0.01-0.38) | 0.18 (0.53-1.60) | 1.12 (0.41-2.22) | 0.9 (0.25-1.78) |
| Day 28 | 0.14 (0.02-0.25) | 0.30 (0.13-0.90) | 0.74 (0.20-1.46) | 0.77 (0.26-1.83) |
| **IFN-γ CD4+/CD8+ T-cell (IU/ml), median (IQR)** | | | | |
| Day 0 | 0.06 (0.01-0.198) | 0.12 (0.03-0.25) | 0.10 (0.02-0.23) | 0.11 (0.00-0.31) |
| Day 14 | 0.12 (0.05-0.43) | 0.88 (0.48-2.32) | 2.22 (0.82-2.70) | 1.68 (0.39-2.68) |
| Day 28 | 0.16 (0.02-0.35) | 0.48 (0.29-1.21) | 1.02 (0.37-2.40) | 1.03 (0.40-2.31) |

**Supplementary Figure 1.** Participant flow chart in the heterologous booster cohorts.

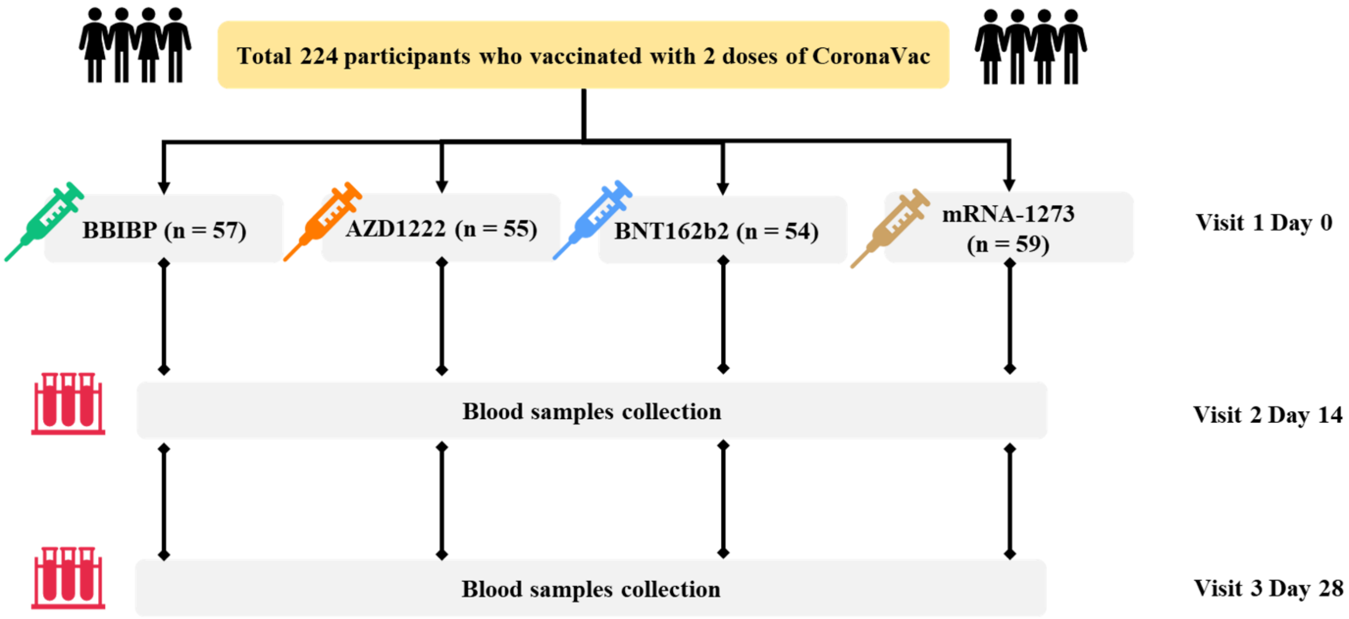

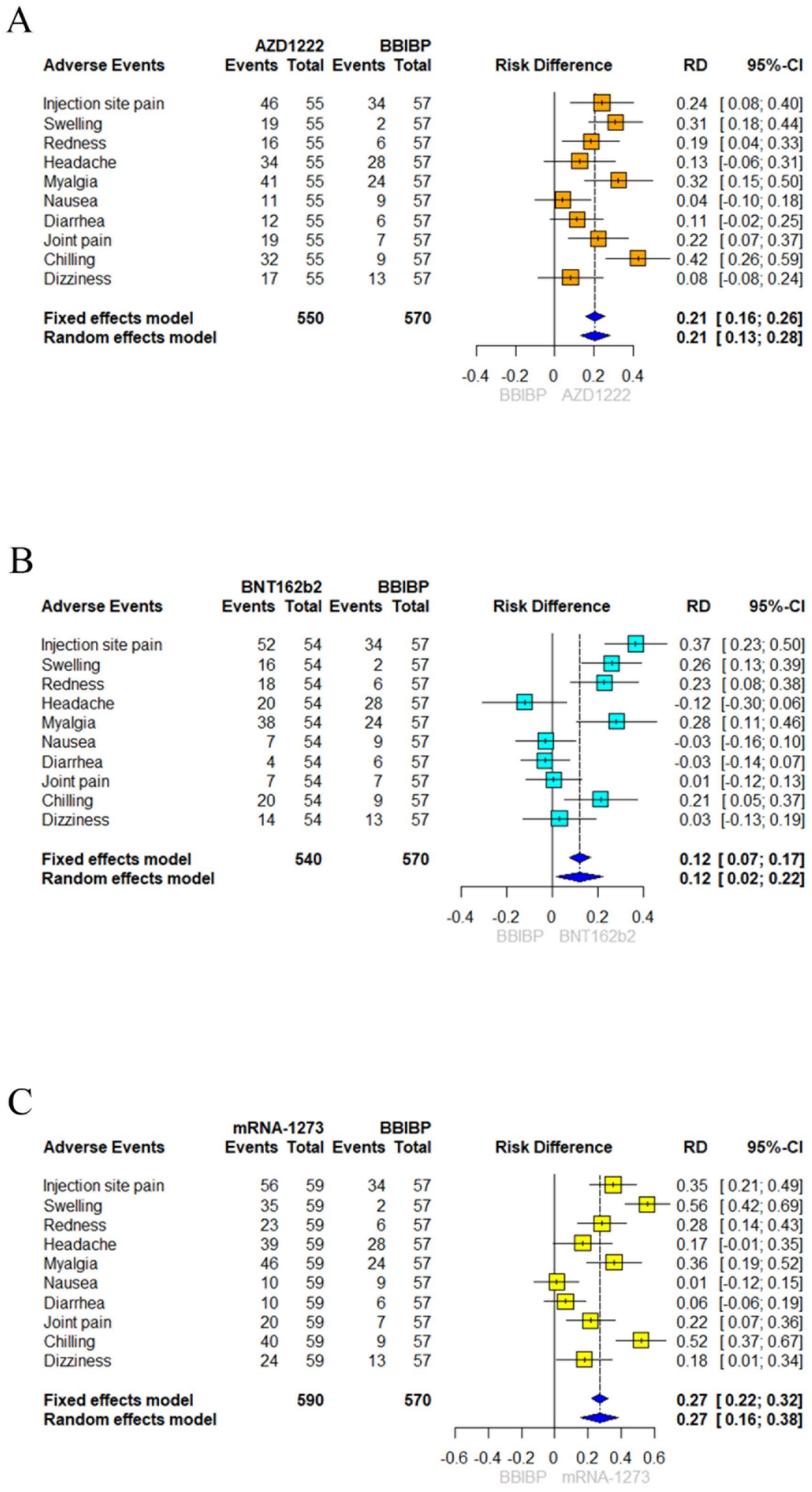


**Supplementary Figure 2.** Forest plot presents any grade the solicited local and systemic adverse events (AEs) across 7 days post-boost vaccination and the absolute risk differences comparison between BBIBP to AZD1222, BNT162b2, and mRNA1273 in the proportion of participants with 95% confidence intervals.

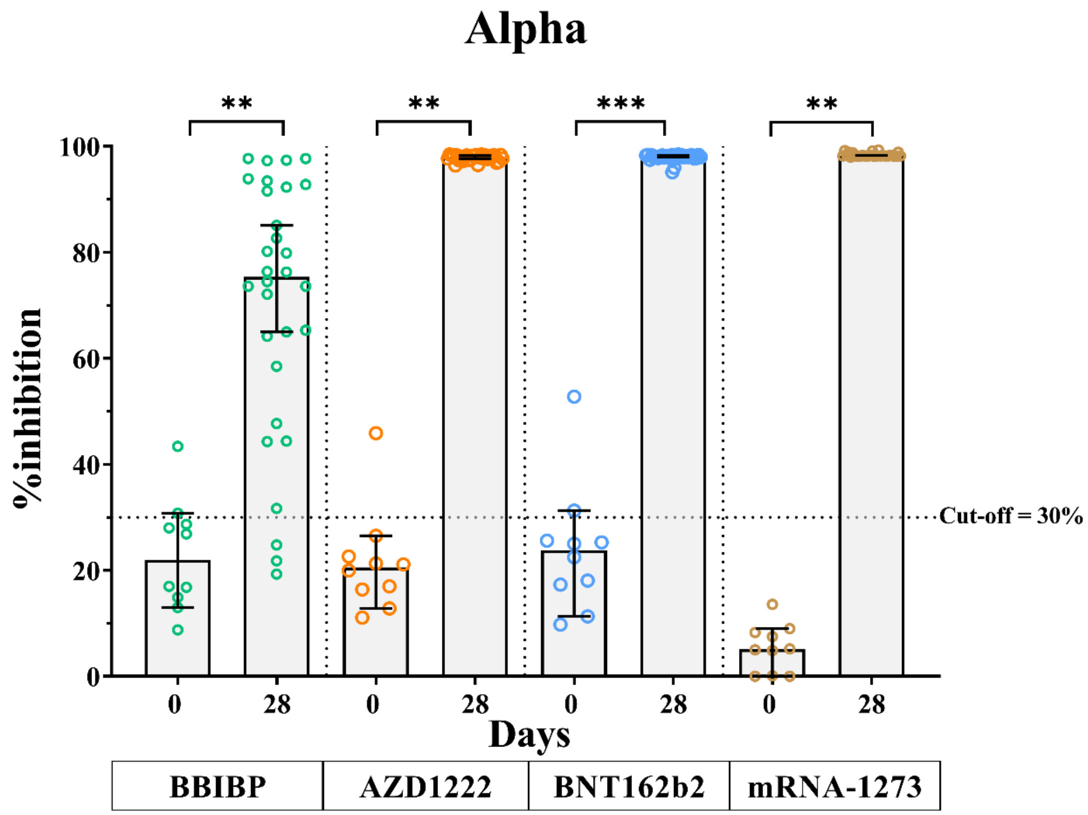


**Supplement Figure 3**. The neutralizing activities against alpha variant. Each data point is represented of the individual who received a heterologous third booster vaccine, the inactivated vaccine, BBIBP (green), the viral-vector vaccine, AZD1222 (orange), the mRNA vaccine, BNT162b2 (blue), or mRNA-1273 (gold). Lines represent median (IQR). ns indicates no significant difference; p <0.01 (**), p <0.001 (***).
